# Feasibility and preliminary effectiveness of behavioral activation for cancer patients with depression

**DOI:** 10.1101/2022.10.06.22280763

**Authors:** Takatoshi Hirayama, Yuko Ogawa, Yuko Yanai, Akie Shindo, Moeko Tanaka, Shin-ichi Suzuki

**Author notes:** **<u>Corresponding author</u>** Takatoshi Hirayama, MD, Department of Psycho-Oncology, National Cancer Center Hospital, 5-1-1 Tsukiji, Chuo-ku, Tokyo, 104-0045 Japan.

## Abstract

**Background:** Behavioral activation (BA) is a psychotherapy that directly approaches patients’ most valued daily activities. BA may be particularly useful for treating depression in patients with cancer, but there is insufficient evidence in this population. This study aimed to verify the feasibility and preliminary effectiveness of BA for patients with depression with all types and stages of cancer.

**Methods:** This pre–post study without a control group was conducted in patients with cancer and depression. The program completion rate was compared with those of previous studies to verify feasibility. To examine the preliminary effectiveness of BA, outcomes were evaluated four times: before and immediately after the program, and 2 weeks and 3 months after the program ended. The primary outcome was the remission rate of depression determined using a binomial test and the 17-item version of the GRID Hamilton Rating Scale for Depression (HAMD_17_). Secondary outcomes were self-reported depression, anxiety, quality of life, changes in behavior, values, and perceived reward of activity and environmental factors. Pre- and post-program data were compared using paired-samples t-tests, and data obtained at four time points were analyzed using one-way repeated measures analysis of variance. Hedge’s g was computed for calculating effect sizes.

**Results:** Participants were recruited from February 2018 to January 2022. Of the 68 patients who were initially recruited, 32 were registered. The completion rate was 75% (24/32), which was similar to previous studies. The total HAMD_17_ score significantly improved after the program with large effect sizes (Hedge’s g =1.95). The remission rate of depression was 62.5% (20/32), which was above the defined threshold value (30%). All but two secondary outcomes significantly improved after the program (p<0.05).

**Conclusions:** This study suggested the feasibility and preliminary effectiveness of BA for patients with depression. To establish further evidence for the use of BA in patients with cancer, randomized controlled trials are needed.

**Trial Registration:** University Hospital Medical Information Network (UMIN CTR) UMIN000036104. Registered 6 March 2019 - Retrospectively registered, https://center6.umin.ac.jp/cgi-open-bin/ctr_e/ctr_view.cgi?recptno=R000041129

## Background

In the oncology setting, psychiatric disorders such as major depression, anxiety disorder, and adjustment disorders are reported to occur in 30–40% of patients [1] and lead to adverse outcomes such as deterioration of quality of life (QOL), lower adherence to chemotherapy, longer hospitalization, and increased suicide risk [2-5]. Appropriate treatment of psychiatric disorders in this population is crucial.

According to international guidelines [6], psychological interventions should be considered as the first-line therapeutic approach for depression in patients with a chronic physical health problem, including cancer, and pharmacological treatment should be added for more severe depression [7]. A meta-analysis reported that psychotherapy was effective for patients with cancer and depression [8]. Therefore, most patients with cancer and depression are candidates for psychotherapy.

There is evidence that patients with cancer may benefit from psychotherapies such as cognitive restructuring–based cognitive behavioral therapy (CBT) and problem-solving therapy [9, 10]. However, since it is common for patients to experience unpleasant thoughts and feelings regarding their experience with cancer [11], quite a few patients with cancer are reluctant to directly face their cancer-related concerns. Furthermore, one common reaction is to attempt to avoid negative feelings. This causes patients to distance themselves from positive aspects of their lives and to lose contact with the very circumstances in which the solutions to their emotional problems can be found [12]. The emotional struggles of patients with cancer appear to be related to behavioral restriction, which reduces exposure to the rewarding and valuable aspects of life [13].

Behavioral activation (BA) may be particularly useful for treating the emotional difficulties of patients with cancer due to its emphasis on eliminating avoidance and encouraging involvement in life-giving activities. BA is a psychotherapy that directly approaches patients’ most valued daily activities. The mechanisms underlying BA are as follows: 1) individuals suffering from depressive or anxiety conditions experience avoidance and decreased participation in normal activities; 2) this leads to decreased opportunities for them to experience joy; 3) they feel that they are overwhelmed by hardships; 4) the value they place on their lives and themselves is diminished; 5) they pay more attention to negative information; 6) they eventually experience more distress and depression; 7) BA facilitates activities they value and breaks this vicious cycle; and 8) this improves the depressive condition and 9) leads to improvement in quality of life. Of note, BA emphasizes identifying values as a part of behavioral change [14]. Thus BA, unlike cognitive restructuring–based CBT, is not a cognitive approach and may therefore be useful for cancer patients who are reluctant to talk about their concerns [15].

The effectiveness of BA for patients with depression is well established [16]. It is as beneficial as CBT in this population, and is relatively inexpensive, costing 21% less than CBT [17]. Because the methods of BA are quite simple, the technique can be learned in a short time by medical staff. By standardizing both methodology and quality, it may be possible to enhance overall cancer treatment quickly, easily, and with low cost.

However, there is insufficient evidence regarding the use of BA in patients with cancer overall, as previous studies have targeted very narrow groups such as patients with breast cancer or cancer-free survivors [18, 19]. In the study by Hopko et al. [18], all subjects were female breast cancer patients, predominantly Caucasian. Only 2 percent of the subjects had stage IV cancer. In the study by Fernández-Rodríguez C, et al. [19], all subjects were cancer-free survivors, predominantly female breast cancer survivors.

Therefore, this study aimed to verify the feasibility and preliminary effectiveness of BA for patients with depression with all types and stages of cancer.

## Methods

### Study design and procedures

This pre–post study without a control group was conducted to examine the feasibility and preliminary effectiveness of BA for patients with cancer and depression. The program completion rate was compared with those of previous studies to verify the feasibility. The primary outcome was the remission rate of depression based on the 17-item version of the GRID Hamilton Rating Scale for Depression (HAMD_17_). Secondary outcomes were self-reported depression, anxiety, QOL, changes in behavior, values, and perceived reward of activity and environmental factors. Outcomes were evaluated four times: before and immediately after the program, and at 2 weeks and 3 months after the program ended.

The study outline and subject eligibility criteria were announced to each department of the National Cancer Center Hospital in Japan through bulletins, in-hospital emails, and websites. Patients who wished to participate in this study, either on their own initiative or on the recommendation of their attending physician, were evaluated in a preliminary interview regarding their eligibility.

This study was approved by the National Cancer Center Institutional Review Board (approval number, 2017-276) and registered at the UMIN Clinical Trials Registry (UMIN000036104). We obtained written consent from all participants.

### Study participants

The eligibility criteria were as follows: patients with cancer who 1) were undergoing cancer treatment or with a history of cancer; 2) met the diagnostic criteria for major depressive episodes according to the Mini-International Neuropsychiatric Interview [20]; 3) had a HAMD_17_ score of ≥8 (depression of mild severity or worse); 4) were aged 20–64 years or aged ≥65 years with a Mini-Mental State Examination (MMSE) [21] of ≥24 points; 5) had an Eastern Cooperative Oncology Group performance status (PS) score of 0–1, as this facilitates behavior modification; 6) could speak Japanese; and 7) provided written consent to participate in this study. This study included patients with all cancer types and stages.

The exclusion criteria were as follows: 1) serious physical or psychiatric symptoms (cognitive dysfunction, impaired consciousness, severe depression wi19th psychiatric symptoms, imminent suicidal ideation, past suicide attempt history) that would prevent program completion; 2) prior intervention by a specialist who conducts BA; and 3) a decision by a researcher or the attending physician of an oncology department that it would be difficult for the individual to participate in the study. For those aged ≥65, or those who did not understand the usual instructions, the MMSE was performed during the preliminary interview, and a score of ≤23 was considered as indicating cognitive dysfunction [21].

While this program did not impact patients’ regular treatment in any way, special psychotherapies such as BA (conducted elsewhere) and CBT were prohibited. We did not limit or adjust medication use because some patients required medications in conjunction with psychotherapy for the treatment of depression.

### Sample size

The goal of depression treatment is not only to reduce symptoms but also to achieve remission, which is defined as the near resolution of symptoms [22]. The remission rate threshold in the current study was 30% and the expected remission rate was 55%, based on the remission rates of 23% (13/56) and 56% (14/25) in the usual treatment group and BA group, respectively, in a study of BA in patients with depression in which HAMD_17_ was used as the endpoint [23]. In the evaluation of the remission rate based on the HAMD_17_, if the post-program rate was at least 30%, the program was judged to be useful.

Twenty-five cases were required based on a binomial test that assumed the following: remission rate threshold = 30%, expected remission rate = 0.55, α = 0.05 (one side), and 1 − β = 0.80. Assuming that 20% of the cases would drop out (i.e., would not complete all seven sessions), we set the sample size at 32 cases.

### BA program

The program consisted of seven 50-min sessions conducted over 1 or 2 weeks, with an average of 5–10 minutes of homework per day. The themes and contents of the program are shown in Table 1.

**Table 1.**
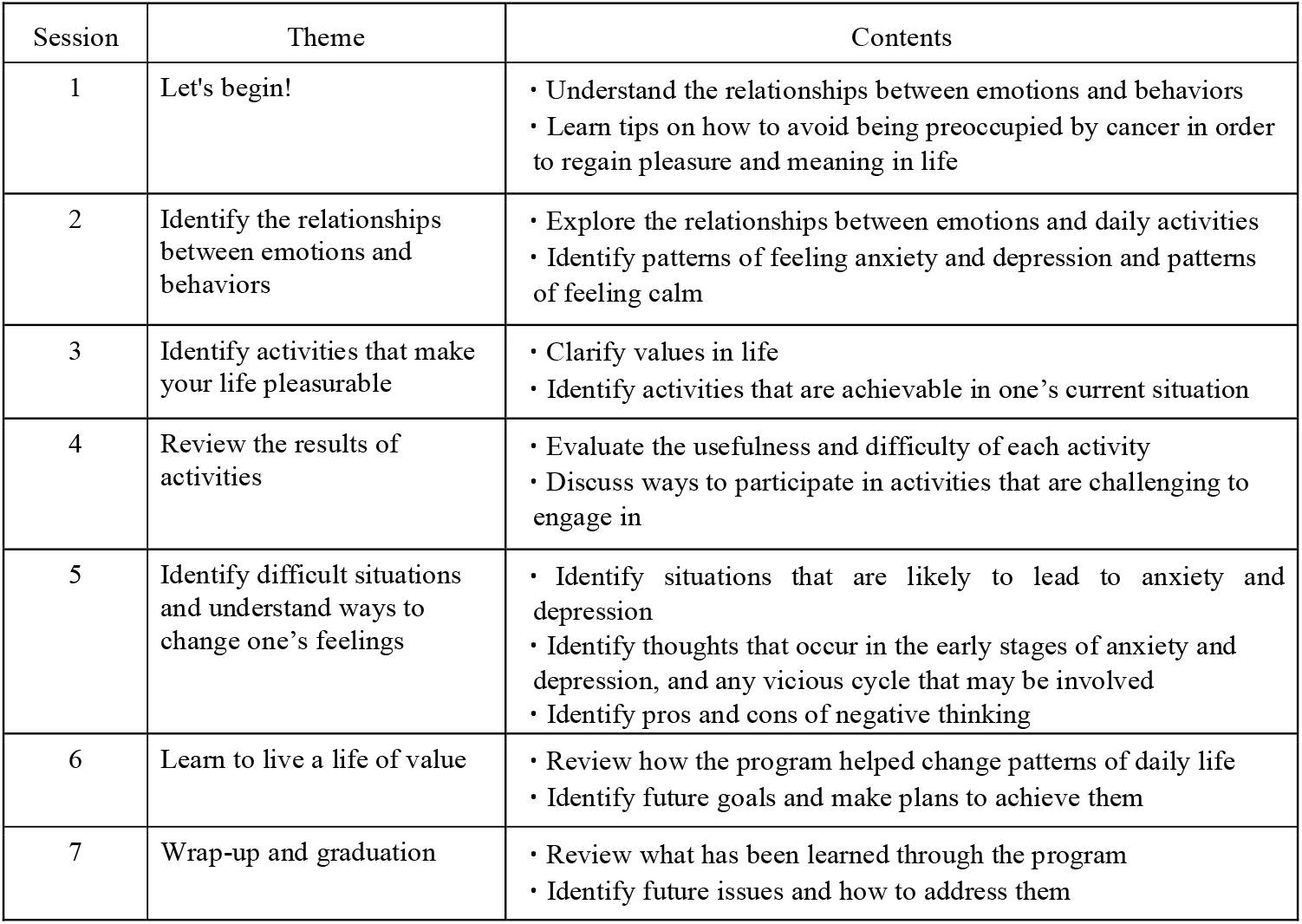
Themes and contents of the behavioral activation program

The program was verified in a pilot study that explored the applicability of BA to patients with cancer [15]. The results showed that the program completion rate and remission rate of depressive symptoms were high regardless of cancer stage.

All therapists were required to be psychotherapists or psychiatrists with clinical experience including work with patients with cancer, and to have sufficient experience conducting BA either in the pilot study [15] or in BA training. Trainees observed seven BA sessions conducted in a clinical setting by a supervisor with sufficient experience with BA. After observing these sessions, the trainees conducted the BA on their own, albeit under supervision. Thereafter, the trainees were certified as having sufficient experience to conduct BA.

### Measures

The program completion rate (the number of subjects who completed all seven sessions divided by the number of subjects who participated in the program, multiplied by 100) was calculated to verify feasibility.

The primary outcome was the remission rate among patients with depression, as determined by the HAMD_17_. Secondary outcomes were the Beck Depression Inventory-II (BDI-II), Generalized Anxiety Disorder-7 (GAD-7), Functional Assessment of Cancer Therapy-General (FACT-G), Behavioral Activation for Depression Scale-Short Form (BADS-SF), Valuing Questionnaire (VQ), and Reward Probability Index (RPI). The individuals responsible for measuring outcomes and assessing patients differed from those who were responsible for conducting the program.

The HAMD_17_ is an amended version of the original Hamilton Depression Rating Scale that provides standardized explicit scoring conventions with a structured interview guide for administration and scoring [24, 25]. The severity of major depressive symptoms was assessed using the HAMD_17_ score, as follows: 0–7, no depression; 8–16, mild depression; 17–23, moderate depression; and ≥24, severe depression [26].

The BDI-II is used to measure depressive symptoms and consists of 21 self-reported items scored on a 4-point scale [27]. The psychometric criteria for the BDI-II are generally considered to be excellent when the instrument is administered to outpatients [27, 28]. Good reliability and validity have been reported for the Japanese version [29].

The GAD-7 is a seven-item questionnaire developed to identify probable cases of GAD and to measure the severity of GAD symptoms [30]; a Japanese version has been validated [31]. The total score of the GAD-7 ranges from 0 to 21.

The FACT-G was used to evaluate QOL [32]. This widely used questionnaire consists of 27 items, with a higher score indicating better QOL. The questionnaire comprises four domains: physical, social, emotional, and functional well-being.

The BADS-SF was developed to assess changes in behavior resulting from BA [33]. The BADS-SF comprises subscales regarding two traits, Activation and Avoidance. The Japanese version of this scale consists of eight items, which is one fewer than in the original version, and the validity and reliability have been confirmed [34].

The VQ is a 10-item self-reported questionnaire that measures the degree to which one’s behaviors are consistent with their values [35]. The score of each item in the VQ was calculated for each of the two factors, VQ Progress (VQ-P) and VQ Obstruction (VQ-O). The VQ-P measures the extent to which individuals are aware of what is personally important to them and their perseverance in achieving whatever this is. The VQ-O measures the extent to which living in line with one’s values is disrupted by avoiding experiences that distract from this goal, due either to neglect or to a focus on other psychological experiences. The validity and reliability of the Japanese version have been confirmed [36].

The RPI was used to assess environmental reward [37]. The Japanese version of the RPI consists of 19 items (one fewer than the original scale) across three factors: Amount of Reward, Environmental Suppressors, and Reward Skill. The validity and reliability of the Japanese version have been confirmed [38].

### Data analysis

#### Feasibility

In two previous studies of BA for patients with cancer, the completion rates were 76.2% (32/42) [18] and 77.3% (17/22) [19], respectively. In this study, BA was considered to be feasible if the completion rate was at least 75%. The completion rate of the program was calculated using the number of participants in the program as the denominator and the number of people who completed all seven sessions as the numerator.

#### Major statistical analysis

To clarify the preliminary effectiveness of BA for depression in patients with cancer, the HAMD_17_-based remission rate was determined as follows: (the number of patients with a HAMD_17_ score of ≤7 points at the end of the program) / (the number of patients who participated in the program [including dropouts]). The remission rate was evaluated by a binomial test; if the remission rate after program completion was at least 30%, which was the value defined as the threshold, BA was judged to be useful.

All assessors received extensive HAMD_17_ training. The reliability of the interview ratings was determined by 13 random samples (10.2%), and the interrater agreement (kappa) value for the diagnosis of depression was 1.0, indicating excellent interrater reliability.

#### Secondary statistical analysis

Exploratory secondary statistical analysis was performed to supplement the main statistical analysis results. Quantitative data were analyzed using IBM SPSS Statistics version 28 (IBM Corp, Armonk, NY, USA). The significance level was 5% on both sides. For the BDI-II and GAD-7, the total score of each scale was calculated. For the FACT-G, BADS-SF, VQ, and RPI, each subscale and total score were calculated.

For parametric data, comparison of data obtained before and immediately after the end of the program (two time points) were analyzed using paired-samples t-tests, and data at four time points (before and immediately after the program, and 2 weeks and 3 months after the program ended) were analyzed using one-way repeated measures analysis of variance. For nonparametric data, the Wilcoxon signed-rank test was used to compare pre- and post-program responses.

To ease interpretation, Hedge’s g was computed, such that values of 0.2, 0.5, and 0.8 denoted small, moderate, and large effect sizes, respectively [39].

## Results

### Participants

Participants were recruited from February 2018 to January 2022. Of the 68 patients who were initially recruited, 32 were registered in this study, and 24 completed it (completion rate, 75% (24/32)) (Fig. 1). Thirty-six patients were not registered because they did not meet the diagnostic criteria for major depressive episodes according to the Mini-International Neuropsychiatric Interview.

**Figure.1.**
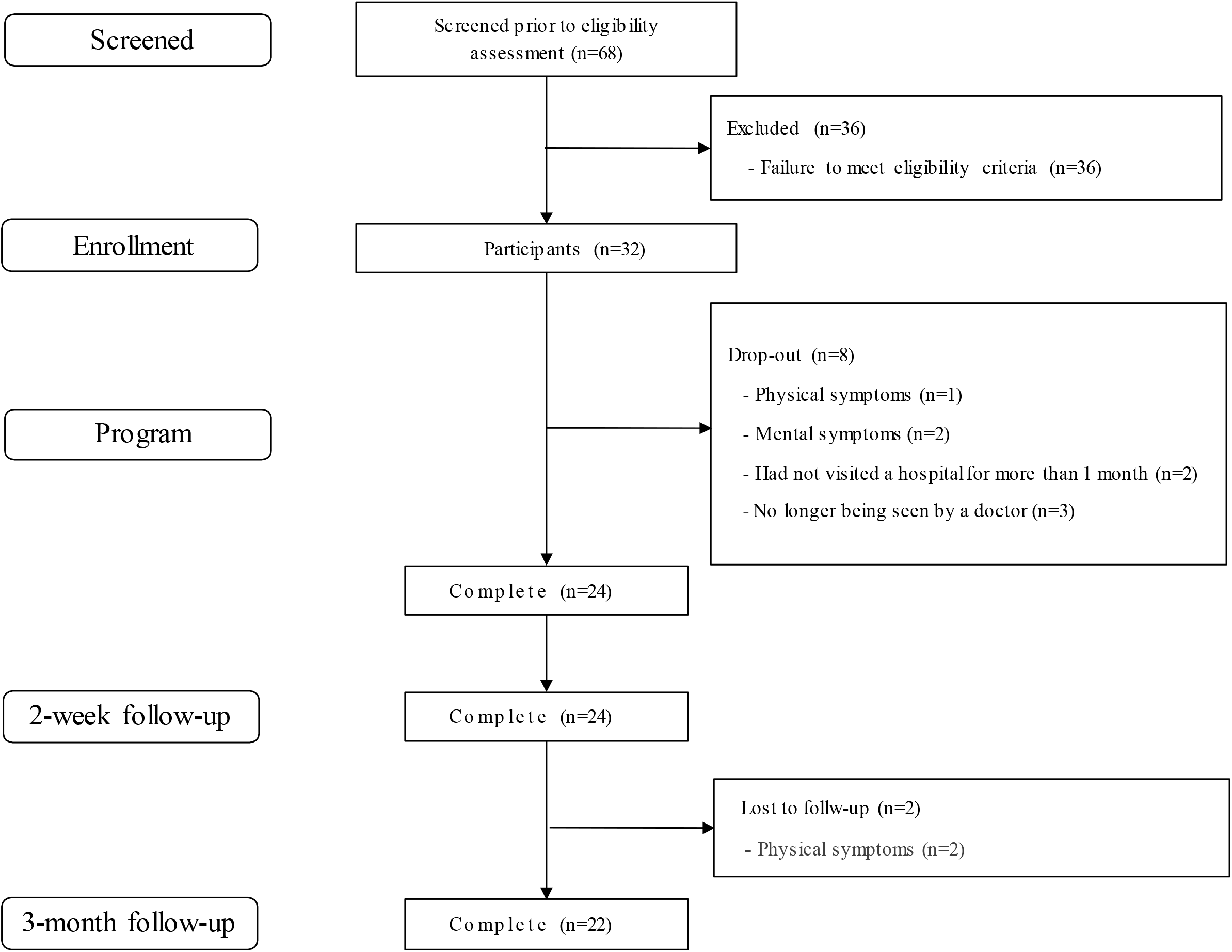
Consolidated Standards of Reporting Trials (CONSORT) Diagram

The demographic characteristics of patients who completed this program are shown in Table 2. Eighteen females and six males with an average age of 55.9 years (SD 10.0) were included. Cancer types included breast cancer (n=10, 41.7%), lung cancer (n=2, 8.3%), colon cancer (n=2, 8.3%), retroperitoneal tumor (n=2, 8.3%), and other (n=8, 33.3%). The most common stage at diagnosis was stage 4 (n=10, 41.7%), followed by stage 1 (n=8, 33.3).

**Table 2.**
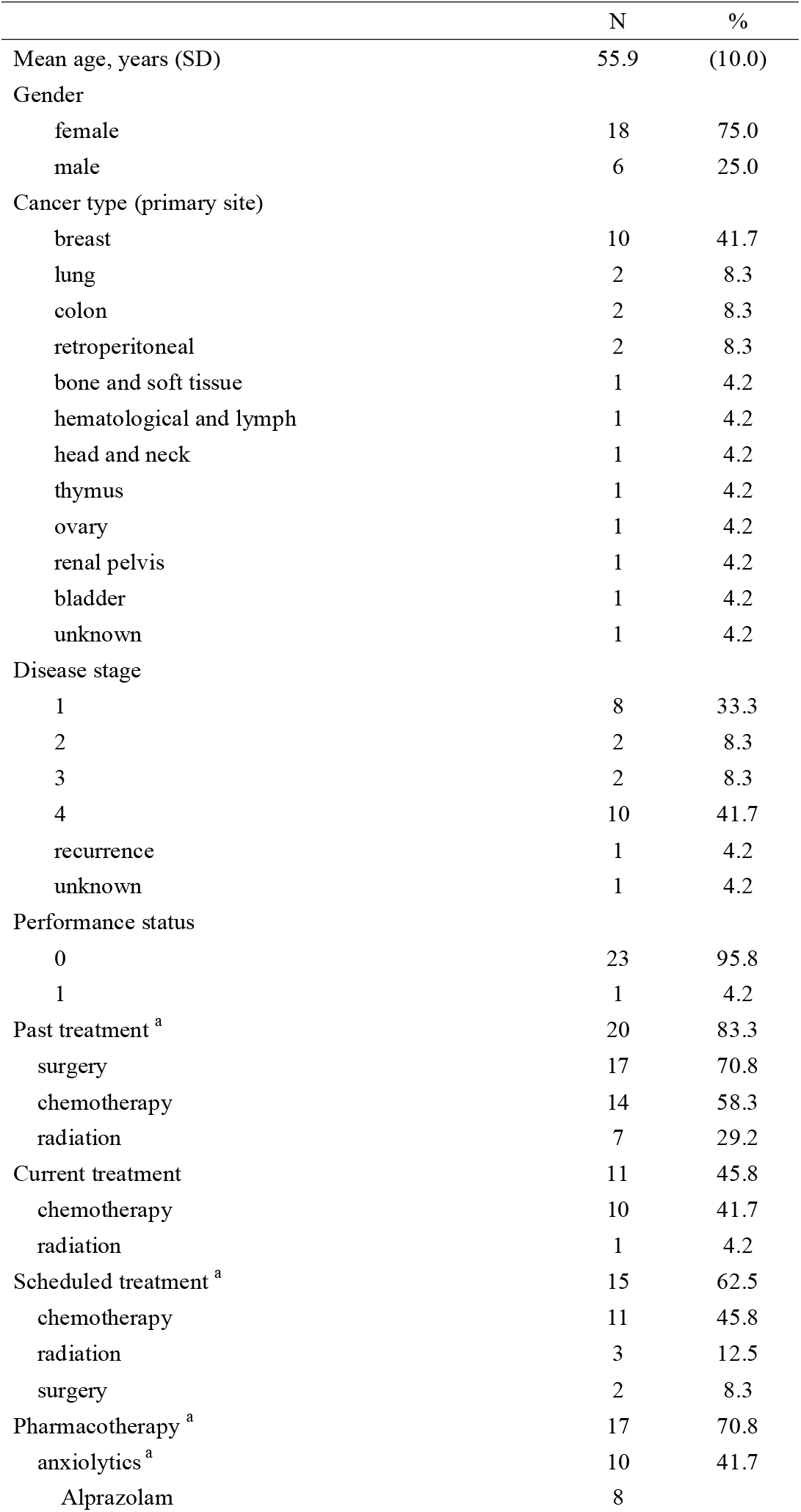
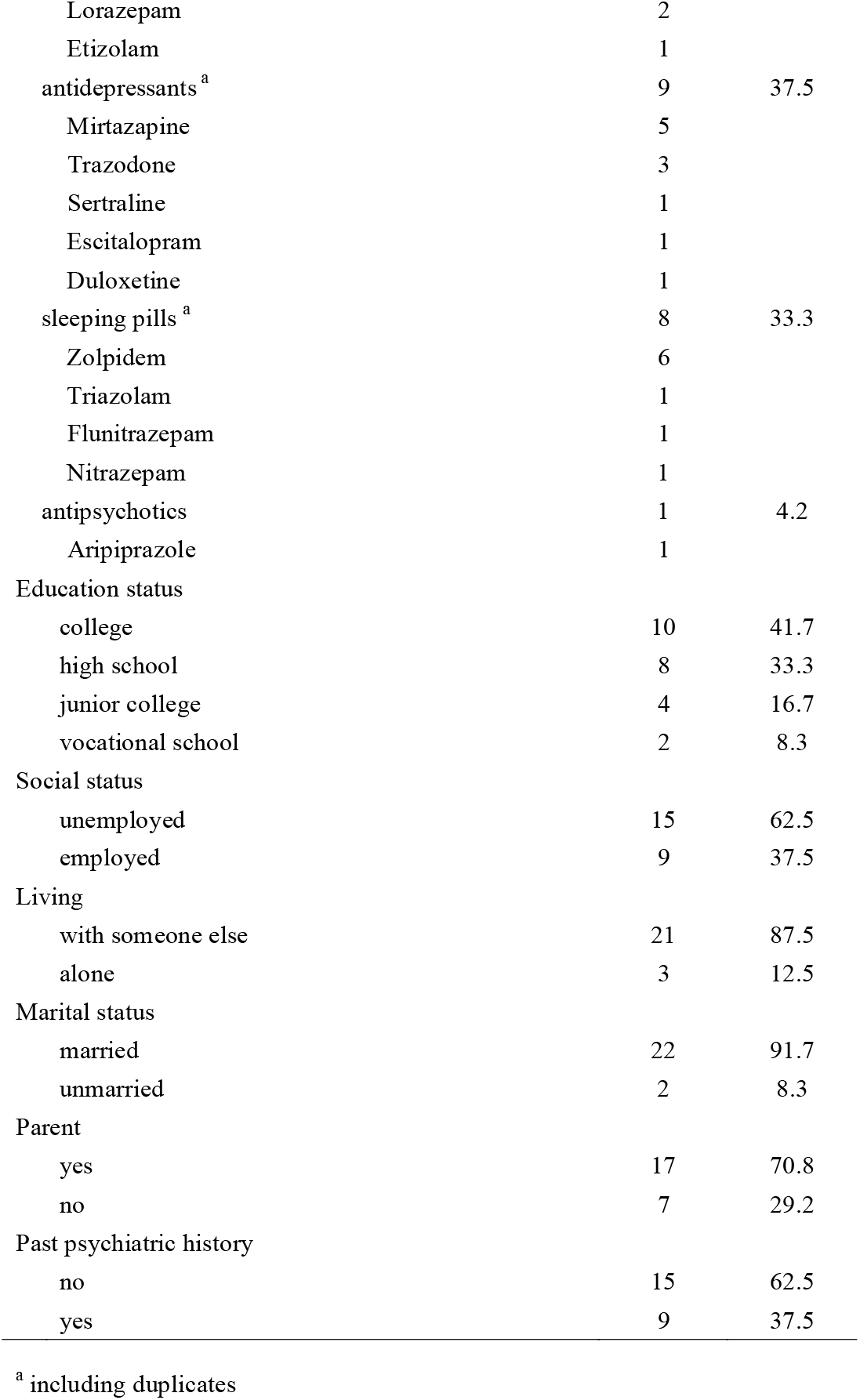
Demographic characteristics of patients who completed the program

### Primary Outcome

The total HAMD_17_ score significantly improved after the program with large effect sizes (Hedge’s g =1.95) (Table 3). The remission rate of depression was 62.5% (20/32). BA was judged to be useful because the remission rate after the program was greater than the pre-defined threshold of 30%.

**Table 3-1.** Score differences before and after the program (paired t-test, two-sided test)

|  | Pre |  | Post |  | *t* | *p* | *Hedge's g* |
| --- | --- | --- | --- | --- | --- | --- | --- |
|  | *M* | *SD* | *M* | *SD* |  |  |  |
| HAMD<sub>17</sub> | 17.9 | 5.2 | 4.4 | 4.9 | 9.9 | <0.001 | 1.95 |
| BDI-II | 29.0 | 9.0 | 16.0 | 10.9 | 5.4 |  | 1.06 |
| GAD-7 | 11.9 | 5.3 | 6.1 | 4.9 | 6.2 |  | 1.23 |
| FACT-G |  |  |  |  |  |  |  |
| Physical | 13.3 | 6.2 | 17.1 | 8.1 | -2.9 | 0.008 | -0.58 |
| Social/Family | 13.0 | 6.4 | 16.0 | 6.2 | -3.2 | 0.004 | -0.63 |
| Emotional | 8.5 | 5.0 | 13.2 | 5.5 | -5.0 | <0.001 | -0.98 |
| Functional | 8.5 | 3.6 | 13.9 | 5.4 | -5.9 |  | -1.17 |
| Total | 43.3 | 12.7 | 60.3 | 19.3 | -5.4 |  | -1.06 |
| VQ Obstruction | 15.1 | 4.4 | 14.5 | 5.2 | 0.6 | 0.575 | 0.11 |
| BADS-SF |  |  |  |  |  |  |  |
| Activation | 6.2 | 3.4 | 11.9 | 5.0 | -6.1 | <0.001 | -1.19 |
| Avoidance <sup>a</sup> | 12.7 | 3.4 | 15.0 | 3.4 | -3.8 |  | -0.75 |
| Total | 18.9 | 4.9 | 27.0 | 6.4 | -6.2 |  | -1.23 |
| RPI Environmental Suppressors <sup>a</sup> | 22.5 | 3.8 | 24.8 | 3.7 | -2.8 | 0.010 | -0.55 |

**Table 3-2.**
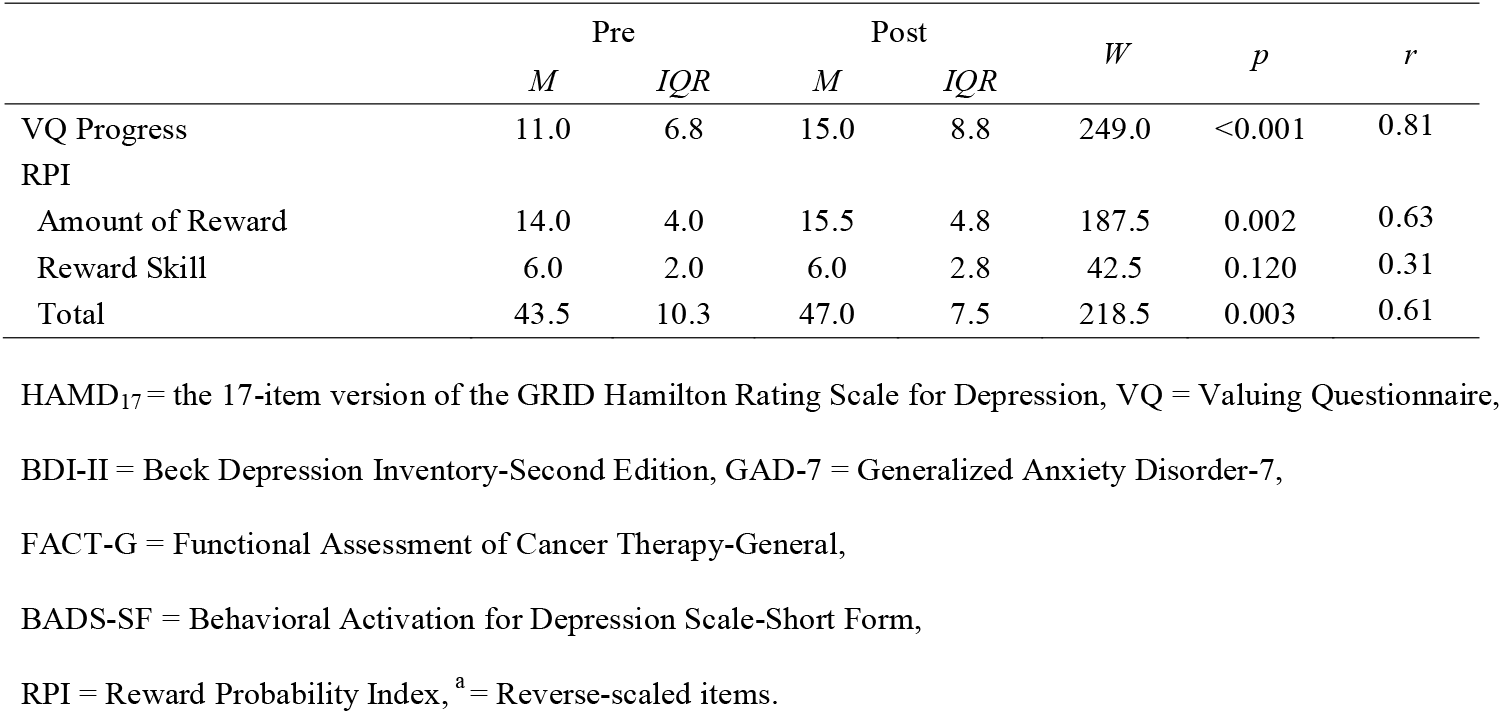
Score differences before and after the program (Wilcoxon signed-rank test, two-sided test)

|  | Pre |  | Post |  | *W* | *p* | *r* |
| --- | --- | --- | --- | --- | --- | --- | --- |
|  | *M* | *IQR* | *M* | *IQR* |  |  |  |
| VQ Progress | 11.0 | 6.8 | 15.0 | 8.8 | 249.0 | <0.001 | 0.81 |
| RPI |  |  |  |  |  |  |  |
| Amount of Reward | 14.0 | 4.0 | 15.5 | 4.8 | 187.5 | 0.002 | 0.63 |
| Reward Skill | 6.0 | 2.0 | 6.0 | 2.8 | 42.5 | 0.120 | 0.31 |
| Total | 43.5 | 10.3 | 47.0 | 7.5 | 218.5 | 0.003 | 0.61 |

HAMD<sub>17</sub> = the 17-item version of the GRID Hamilton Rating Scale for Depression, VQ = Valuing Questionnaire,
BDI-II = Beck Depression Inventory-Second Edition, GAD-7 = Generalized Anxiety Disorder-7,
FACT-G = Functional Assessment of Cancer Therapy-General,
BADS-SF = Behavioral Activation for Depression Scale-Short Form,
RPI = Reward Probability Index, <sup>a</sup> = Reverse-scaled items.

### Secondary Outcomes

Regarding changes before and after the program, normality was confirmed for all outcomes except VQ-P, RPI Amount of Reward, RPI Reward Skill, and RPI Total.

All secondary outcomes except VQ-O and RPI Reward Skill were significantly improved immediately after the program (p<0.05) (Table 3) and 3 months later (Table 4). No adverse events were observed during the study.

**Table 4-1.** One-way repeated measures ANOVA

|  | Pre |  | Post |  | 2-week follow-up |  | 3-month follow-up |  | *F* | *p* | *partial*<br>*η*<sup>2</sup> | Multiple Comparison |
| --- | --- | --- | --- | --- | --- | --- | --- | --- | --- | --- | --- | --- |
|  | *M* | *SD* | *M* | *SD* | *M* | *SD* | *M* | *SD* |  |  |  |  |
| HAMD<sub>17</sub> | 17.5 | 5.2 | 4.1 | 4.5 | 5.0 | 5.0 | 5.9 | 7.3 | 46.50 |  | 0.69 |  |
| BDI-II | 29.1 | 9.3 | 15.9 | 11.4 | 15.7 | 11.4 | 14.6 | 12.2 | 24.02 | <0.001 | 0.53 | Pre > Post, 2-week, 3-month |
| GAD-7 | 12.2 | 5.3 | 6.4 | 5.0 | 6.8 | 5.3 | 6.5 | 5.4 | 22.11 |  | 0.51 |  |
| FACT-G |  |  |  |  |  |  |  |  |  |  |  |  |
| Physical | 13.1 | 6.2 | 17.4 | 8.4 | 18.0 | 7.8 | 18.9 | 7.9 | 10.91 | <0.001 | 0.34 | Pre < Post, 2-week, 3-month |
| Social/Family | 13.2 | 6.6 | 16.2 | 6.5 | 13.7 | 7.7 | 15.9 | 6.8 | 3.75 | 0.015 | 0.15 | Pre < Post, 3-month |
| Emotional | 8.3 | 5.1 | 13.1 | 5.7 | 13.8 | 5.5 | 13.5 | 5.3 | 18.98 |  | 0.48 |  |
| Functional | 8.7 | 3.6 | 14.5 | 5.2 | 13.8 | 4.8 | 15.4 | 5.9 | 24.18 | <0.001 | 0.54 | Pre < Post, 2-week, 3-month |
| Total | 43.2 | 13.1 | 61.1 | 19.8 | 59.2 | 19.8 | 63.7 | 20.8 | 20.31 |  | 0.49 |  |
| VQ Obstruction | 15.3 | 4.4 | 15.0 | 5.1 | 14.6 | 4.8 | 14.3 | 5.2 | 0.42 | 0.738 | 0.02 | — |
| BADS-SF |  |  |  |  |  |  |  |  |  |  |  |  |
| Activation | 6.4 | 3.3 | 12.3 | 5.1 | 12.0 | 5.7 | 11.7 | 6.1 | 12.13 |  | 0.37 |  |
| Avoidance <sup>a</sup> | 12.7 | 3.4 | 14.9 | 3.5 | 14.2 | 3.4 | 14.6 | 3.1 | 6.45 | <0.001 | 0.24 | Pre < Post, 2-week, 3-month |
| Total | 19.1 | 5.1 | 27.2 | 6.6 | 26.2 | 7.8 | 26.3 | 7.7 | 15.54 |  | 0.43 |  |
| RPI Environmental Suppressors <sup>a</sup> | 22.5 | 4.0 | 24.6 | 3.8 | 25.1 | 4.2 | 25.9 | 4.0 | 6.94 |  | 0.25 |  |

**Table 4-2.** Friedman repeated measures ANOVA on ranks

| | Pre | | Post | | 2-week follow-up | | 3-month follow-up | | $\chi^2$ | $p$ | $r$ | Multiple Comparison |
| --- | --- | --- | --- | --- | --- | --- | --- | --- | --- | --- | --- | --- |
| | $M$ | $IQR$ | $M$ | $IQR$ | $M$ | $IQR$ | $M$ | $IQR$ | | | | |
| VQ Progress | 11.0 | 7.5 | 15.0 | 9.0 | 16.5 | 7.5 | 14.0 | 5.8 | 31.41 | <0.001 | 0.81<br>0.77<br>0.86 | Pre > Post<br>Pre > 2-week<br>Pre > 3-month |
| RPI |  |  |  |  |  |  |  |  |  |  |  |  |
| Amount of Reward | 14.0 | 4.5 | 16.0 | 5.5 | 16.5 | 5.0 | 17.0 | 4.0 | 14.3 | 0.002 | 0.63 | Pre > 3-month |
| Reward Skill | 6.0 | 2.0 | 6.0 | 3.0 | 6.0 | 2.0 | 6.0 | 2.5 | 2.79 | 0.436 | — | — |
| Total | 43.5 | 11.5 | 47.0 | 8.5 | 49.0 | 9.3 | 47.0 | 9.8 | 15.64 | <0.001 | 0.65<br>0.67 | Pre > 2-week<br>Pre > 3-month |

ANOVA = Analysis of variance, HRSD<sub>17</sub> = the 17-item version of the GRID Hamilton Rating Scale for Depression, VQ = Valuing Questionnaire,
BDI-II = Beck Depression Inventory-Second Edition, GAD-7 = Generalized Anxiety Disorder-7, FACT-G = Functional Assessment of Cancer Therapy-General,
BADS-SF = Behavioral Activation for Depression Scale-Short Form, RPI = Reward Probability Index, <sup>a</sup> = Reverse-scaled items.

## Discussion

This intervention study is the first to verify the feasibility and preliminary effectiveness of BA for patients with depression with all types and stages of cancer. The program completion rate was 75% (24/32). This result is consistent with two previous studies in patients with cancer, in which the completion rates were 76.2% (32/42) and 77.3% (17/22) [18, 19], respectively. Together these findings suggest that BA might be feasible for use in patients with cancer. More than half of the initially recruited patients were not registered because they did not meet the diagnostic criteria for major depressive episodes. Recent survey in Japan reported that 9.0% of patients with cancer had depressive symptoms [40]. This low prevalence of depression might have contributed to the low registration rate. Though the prevalence of depression is low in patients with cancer in Japan, we believe our findings are significant because psychiatric disorders including depression lead to various adverse outcomes in this population [2-5].

Because the remission rate of depression was 62.5% (20/32) (≥30%), this program was judged to be useful. This remission rate is similar to that of 56% achieved by Dimidjian et al. in a study of BA for patients with depression in which HAMD_17_ was used as the endpoint [23]. On the other hand, Hopko et al. reported that the HAMD_17_-based remission rate in patients with breast cancer was 71% [18]. Both the current study and that of Hopko et al. had similar HAMD_17_ scores before the program (17.9 (5.2) and 19.2 (7.0), respectively). The discrepancy in remission rates may be due to differences between populations regarding cancer stages; for instance, 41.7% of patients in this study had stage 4 disease, compared to 1% of patients in the study by Hopko et al. Overall, our findings suggest that BA might also be effective in patients with advanced cancer.

All secondary outcomes except VQ-O and RPI Reward Skill were significantly improved after the program (p<0.05). VQ-P measures the extent to which individuals are aware of what is personally important to them and their perseverance in achieving it. BA emphasizes identifying values as a part of behavioral change [14], and this may improve VQ-P rather than VQ-O. RPI Reward Skill measures the skills required to earn rewards that lead to proper operant behavior. Our findings suggested that BA did not affect RPI Reward Skill. However, it is less important to have the skills to earn rewards than it is to use these skills, take actions, and expose oneself to rewards [38].

In previous randomized studies of BA for patients with cancer, BDI-II and BADS were significantly improved after the intervention [18, 19]. Although the scales used in this study were different, BA improved anxiety, health-related QOL, and positive or pleasurable outcomes or rewards that follow behaviors. Our findings are consistent with these results, and BA may not only improve depression in patients with cancer, but may also provide broad support to help patients better deal with the cancer trajectory and lead a more fulfilling life based on their values. This suggests that BA may supplement psychotherapy by serving as a comprehensive support method for patients with cancer.

This study has several limitations. First, it used a single-arm pre–post design without a control arm, which may have caused several systematic biases. To verify the effectiveness of BA, a randomized controlled trial is needed as a next step. Second, this study was conducted at a single cancer center, raising the question of institutional bias, and the results may not be applicable to other settings. Third, the subjects of this study were all Japanese. This is the first study to suggest the feasibility and preliminary effectiveness of BA for Japanese patients with cancer. But, careful consideration should be required when adapting our methodology to other races and ethnic groups. Fourth, although this study included patients with all cancer types, nearly half of them had breast cancer, and there were at most two cases of every other cancer type. The conclusion that BA was preliminary effective for all patients with cancer should be evaluated carefully. Fifth, due to dropouts, this study did not reach the target number of registrations and the results should therefore be interpreted with caution. Finally, the preliminary effectiveness of BA should not be exaggerated due to the possibility of bias, for instance due to the effect of pharmacotherapies and other co-interventions (support from medical staff and their family members). In this study, about 70% of the participants had pharmacotherapies including antidepressants. The effect of pharmacotherapies should be carefully considered, since a systematic review reported low-certainty evidence for antidepressants compared with placebo in the treatment of depression in patients with cancer [41].

## Conclusions

This study suggested the feasibility and preliminary effectiveness of BA for patients with depression with all types and stages of cancer. To establish further evidence for the use of BA in patients with cancer, randomized controlled trials are needed.

## Data Availability

All data produced in the present work are contained in the manuscript.

## List of abbreviations

BA: behavioral activation
HAMD_17_: 17-item version of the GRID Hamilton Rating Scale for Depression
CBT: cognitive behavioral therapy
MMSE: Mini-Mental State Examination
PS: performance status
BDI-II: Beck Depression Inventory-II
GAD-7: Generalized Anxiety Disorder-7
FACT-G: Functional Assessment of Cancer Therapy-General
BADS-SF: Behavioral Activation for Depression Scale-Short Form
VQ: Valuing Questionnaire
VQ-P: Valuing Questionnaire Progress
VQ-O: Valuing Questionnaire Obstruction
RPI: Reward Probability Index
SD: standard deviation IQR: interquartile range
NCCH: National Cancer Center Hospital

## Declarations

### Ethics approval and consent to participate

This study was approved by the National Cancer Center Institutional Review Board (approval number, 2017-276). Written informed consent was obtained from all study participants.

### Consent for publication

We used our institutional consent form and obtained written consent for publication from all participants.

### Availability of data and materials

All data generated or analyzed during this study are included in this published article.

### Authors’ contributions

TH, YO and SS designed this study. TH, YO, YY, AS, and MT performed the BA program. TH and YO analyzed and interpreted the patient data. TH was a major contributor in writing the manuscript. All authors read and approved the final manuscript.

## Acknowledgements

The authors thank all the members of the palliative care team of the National Cancer Center Hospital (NCCH) in Japan, as well as Tomoko Mizuta (Department of Psycho-Oncology, NCCH) and all patients with cancer who participated in this study.

## Authors’ information

Not applicable

